# Global In-Silico Performance Assessment of the CDC Zika Trioplex Detection System

**DOI:** 10.1101/2025.08.14.25333671

**Authors:** Shella Efire, Mignane Ndiaye, Diamilatou Balde, Marielton Dos Pasos Cunha, Andrea Silverj, Manfred Weidmann, Oumar Faye, Idrissa Dieng

## Abstract

Arboviruses, transmitted by arthropod vectors such as mosquitoes and ticks, represent a significant public health challenge due to their association with high mortality rates and the absence of vaccines or effective treatments. Dengue, Zika, and Chikungunya viruses (DENV, ZIKV, and CHIKV) are of particular concern, often co-circulating and causing outbreaks worldwide. The development of molecular detection tools has been crucial for improving diagnostic accuracy and facilitating early outbreak management. However, many detection tools, including the widely adopted CDC Trioplex RT-PCR assay, have not undergone comprehensive evaluation against global viral sequence diversity. This assay simultaneously detects DENV, ZIKV, and CHIKV, yet its performance across emerging viral strains, particularly in regions like West Africa, remains underexplored. We conducted an *in-silico* analysis of the CDC Trioplex assay’s primers and probes against a global dataset of ZIKV strains. Phylogenetic analysis revealed notable differences between the African and Asian ZIKV genotypes. Further, mismatch analyses demonstrated that the CDC Trioplex primers and probes exhibit varying degrees of mismatches across different ZIKV strains, particularly of African genotypes. These mismatches, especially in key primer and probe regions, can significantly impact PCR efficiency, potentially leading to diagnostic failures. Our findings underscore the need for ongoing surveillance of circulating viral strains and the importance of validating diagnostic assays like the CDC Trioplex in diverse geographic regions. Further experimental research is necessary to determine the practical implications of these mismatches for ZIKV detection accuracy, especially in regions with diverse viral strains such as West Africa.

## Introduction

Arboviruses are viruses transmitted by hematophagous arthropod vectors such as mosquitoes, ticks, and sandflies to their amplifying vertebrate hosts (1). According to the World Health Organization (WHO), vector-borne diseases, including those caused by arboviruses, account for around 17% of infectious diseases and cause approximately 700,000 deaths worldwide every year (2) Accessed 26 July. In most cases, there are no approved vaccines or effective treatments for these diseases, making them a major public health problem. Consequently, rapid diagnosis of these diseases for rapid case management is a real challenge and a major necessity, particularly during epidemics. Among the arboviruses that pose a major threat, Dengue virus (DENV), Zika virus (ZIKV), and Chikungunya virus (CHIKV) have been responsible for numerous epidemics in recent decades (3,4), and often cocirculate locally, underlining the need for effective diagnostic tools for these viruses. In this context, molecular detection of arbovirus infections plays a crucial role in early detection and management of cases, facilitating mitigation efforts and for efficient surveillance. Significant progress has been made in developing molecular detection systems for these viruses in recent years (5–9). However, many of these tools have not undergone comprehensive evaluation, leaving their true diagnostic capabilities uncertain (10). This highlights the urgent need for laboratories to adopt standardized testing methods. Additionally, most current detection systems are limited to identifying one virus at a time, a costly and inefficient approach, especially during epidemics where co-circulation of viruses with similar clinical presentations is possible (11,12).

In late 2015, Zika virus spread from the Pacific Islands to South America, triggering a massive outbreak associated with birth defects and neurological complications. In response, the Centers for Disease Control and Prevention (CDC) developed the Trioplex real-time reverse transcriptase polymerase chain reaction (RT-qPCR) test (13) accessed 26 July 2024, designed for the simultaneous detection and differentiation of ZIKV, DENV, and CHIKV infections. This assay, offered free of charge, quickly gained global adoption due to its proven efficacy, and reliability, when tested with samples from Brazil (14).

Despite its widespread use, a comprehensive global evaluation of the assay’s primers across various viral strains has not been published. In our recent study, we observed poor performance of this system in detecting the West African genotype of CHIKV (15). These findings motivated us to apply an in-silico approach to evaluate the potential erosion of ZIKV Trioplex primers and probes when challenged with the global sequence diversity of the virus.

## Materials and Methods

### Genome sequences dataset construction

Complete genomic sequences available for ZIKV (until January 2023) with information about the date and location of isolation were retrieved from the National Center for Biotechnology Information (NCBI) (https://www.ncbi.nlm.nih.gov/genbank/) website in GenBank format to determine the phylogenetic relationships of the sequenced viruses. All sequences were converted to FASTA format. We set an exclusion threshold for sequences that had more than a total of 1% of “N” (https://biopython.org/wiki/Sequence_Cleaner), to avoid duplicated and low-quality sequences with many unknown nucleotides. All taxon labels for the sequences used in the present study are shown in the format: accession number_country of isolation_year of isolation.

### Sequences alignment and Phylogenetic Analysis

The FASTA sequences were aligned using MUSCLE v.3.8.1551 (16). The presence of putative recombinant sequences was assessed and removed using the RDP5 v.5.5 program

(RDP, GENECONV, MaxChi, BootScan, and Siscan) (17) considering the standard settings to avoid spurious adjacency patterns brought by eventual recombinants. A final alignment (dataset-1) of 728 complete polyprotein coding sequences was obtained and inspected using the program AliView v.1.26 (18).

The phylogenetic trees in the ZIKV datasets were inferred using the maximum likelihood (ML) method implemented in IQ-Tree2 version 2.0.7 (19) under GTR model. The robustness of the groupings observed was assessed using an ultrafast bootstrap approximation (UFboot) (20) during 1,000 replicates.

### In Silico analysis

ZIKV CDC trioplex assay primers and probe sequences were individually aligned with the representative ZIKV dataset downloaded in Genbank (step above). The three separated files were individually aligned using MAFFT (21) and then trimmed to retrieve only the portion of the genomes spanning the oligos (primers/probe) binding site. From the obtained trimmed alignment nucleotide changes were called for all individual sequence of the alignment by using the specific oligo sequence as reference. as previously described (22). Called nucleotide changes were stored in a CSV file containing sequences ID, position of nucleotide in the, allele in the reference (Nucleotide in the Oligo), the alternate allele (Nucleotide in the particular sequence position), the length of the oligos. Finally, information’s stored in the CSV file were used to plot the heatmap of mismatch rate of each sequence of the dataset against the particular oligonucleotide of the ZIKV CDC trioplex assay.

### Data Visualizations

Generated CSV files were merged with the downloaded sequences metadata an used for result representation steps. Mean mismatches rate heatmap by viral genotype for each ZIKV CDC trioplex assay and a Choropleth map of the mean mismatch rate per country were generated using R software version 4.4.1 (2024-06-14).

## Results and Discussion

### Phylogenetic analysis of Representative ZIKV sequences circulating globally

A representative dataset of complete ZIKV genome sequences was downloaded from GenBank and used in this study. A total of 728 sequences obtained from samples collected between 1966 to 2021were downloaded. Viral genotype assignment using Genomedetective (https://www.genomedetective.com/app/typingtool/zika/ accessed on 18. August) reveals that the dataset contained 702 (96.42 %) sequences classified as Asian genotype, 24 (3.29

%) as African genotype and surprisingly 1 (0.13 %) sequence from India (OK054351.1) collected in 2021 assigned as ‘related but not part of Asian genotype’. Additionally phylogenetic tree of the different ZIKV sequences was inferred using IQ-Tree2 version 2.0.7 (19). The robustness of the groupings observed was assessed using an ultrafast bootstrap approximation (UFboot) during 1,000 replicates, in order to confirm reported lineages. A bipartite phylogram with a dominant Asian clade and a significantly different African clade also showed a distinct intermediate clade composed of 3 sequences, the most distinct of which showed a identity of only 95.52 % to the closest Asian and of 88.74 % to the closest African genome sequences (Figure S1).

### Global dataset mismatches analysis

### Mismatches count analysis according to Oligo and ZIKV variants

*In silico* analysis revealed mismatches between the primers/probes and their respective binding regions across several viral strains (Figure 2). Stratified by genotype, the mean mismatch rates for the forward primer were 2, 2, and 1 for the African, Asian, and ‘related to Asian genotype’ category, respectively. The reverse primer exhibited a higher mean mismatch rate for the African genotype (3.5), whereas it was nearly zero for the Asian genotype. As for the probe, the Asian genotype showed the highest mean mismatch rate (1.26), while the African genotype sequences demonstrated lower mismatch rates (Figure S2).

Moreover, the number of mismatches within the primer/probe binding regions varied from one to four, depending on the specific primer/probe sequence and viral genotype. ZIKV viral genotype distribution is quite homogeneous with the African genotype confined in Africa (23) while Asian genotype is more widespread world-wide (24). This in addition to the high mismatches rate associated to African genotype against ZIKV CDC trioplex assay oligos was confirmed by the generated country wise mean mismatches rate choropleth map (Figure S3).

For the Asian genotype, most strains showed no mismatches, or fewer than one, in the forward and reverse primers, except for two strains where two mismatches were identified. These findings indicate a relatively high concordance between the CDC Trioplex ZIKV forward and reverse primers and the target regions in Asian genotype sequences used in this study (Figure S3).

However, the probe revealed a significant number of strains with at least one mismatch, including 81 strains that had up to three mismatches (Figure 1). The presence of mutations in the probe have already been reported to affect the ZIKV PCR efficiency of several PCR assays (25–27). The observed primer-template mismatches, combined with probe mismatches, can have a deleterious impact on the amplification and detection of Asian genotype ZIKV strains by the CDC Trioplex. Regarding the African genotype, all strains show at least one nucleotide substitution in the primer binding regions, and only seven strains show no substitutions when compared to the probe. In some cases, a high number of substitutions were found. For example, there are four mismatches in fourteen strains in the ZIKV CDC Trioplex reverse primer. The strain, categorized as ‘related but not belonging to the Asian lineage’ also features five substitutions in both primers and probe. It has been shown that even a single mismatch in the binding region of a primer can notably impact the performance of viral RT-qPCR systems by raising cycle threshold (Ct) values, which reduces their efficiency (28,29). Therefore, having two to four mismatches in the binding sites of the CDC Trioplex ZIKV primers and probe is likely to significantly affect the efficiency of ZIKV detection using the CDC Trioplex. However further experimental research is needed to determine the actual impact of these mutations on the diagnostic efficiency of the CDC Trioplex for ZIKV strains belonging to the African genotype. Nevertheless, it is important for laboratories to be aware of these potential issues.

**Figure 1.**
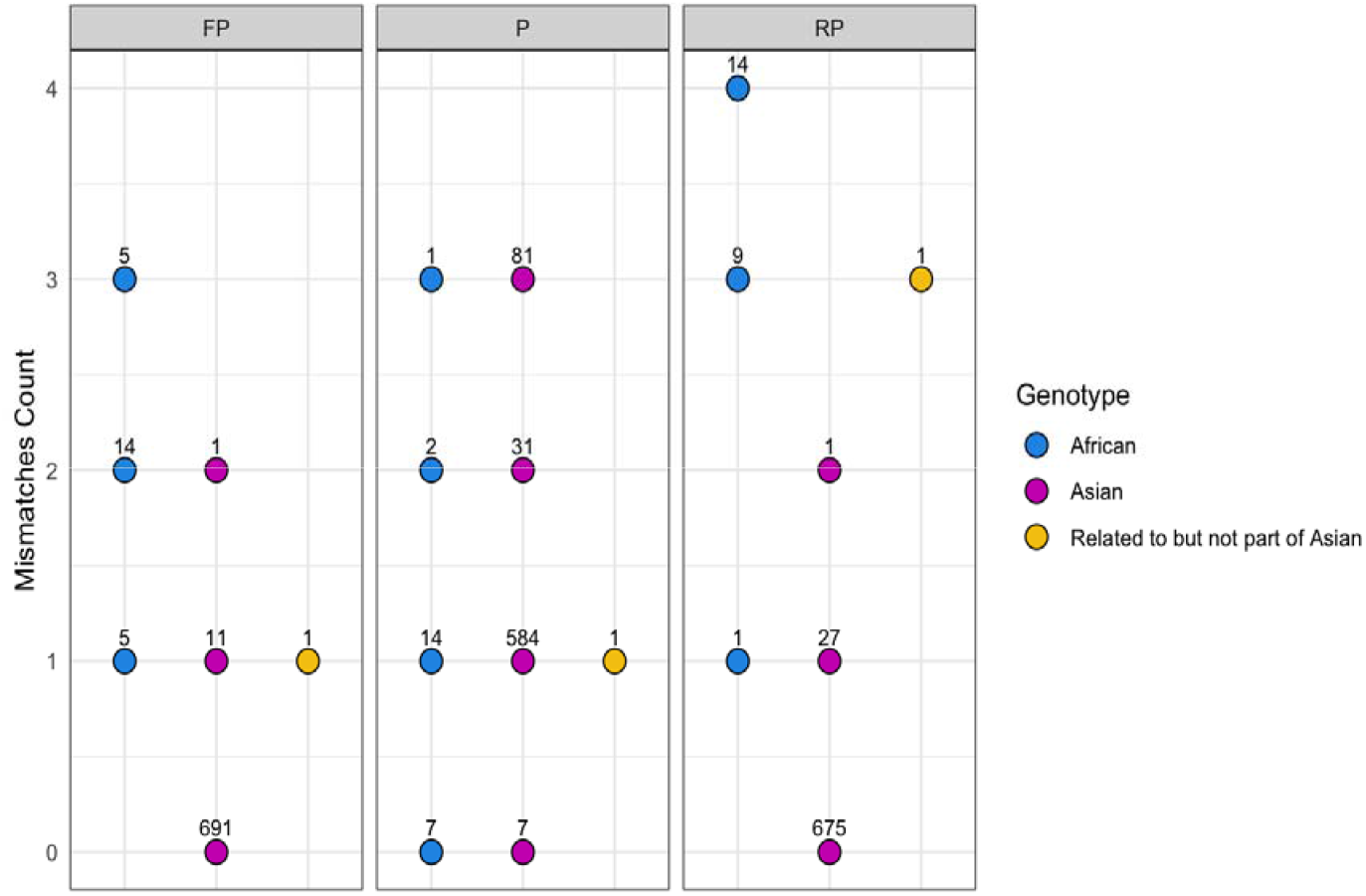
Scatterplot representation of identified mismatches in the ZIKV amplicon of the CDC Trioplex PCR. The y axis represents the number of mismatches, while the x axis the assigned viral Genotype. The number above each dot highlights the number of sequences of a particular genotype exhibiting different numbers of mismatches. forward primer (FP), reverse primer (RP) and probe (P) are represented in individual panels, from left to right.

### MSA mismatches impacts analysis

Using respectively 38, 53, 22 sequence datasets for forward primer, probe, and reverse primer we generated mismatch alignments for the altered primers and probe of the ZIKV amplicon. These mismatches are found in ZIKV strains from several geographical regions, with a notably higher frequency observed in West African strains in the primers (Figure 2). The impact on the efficiency of the RT-qPCR systems can vary based on position of the mismatch (30,31).

**Figure 2.**
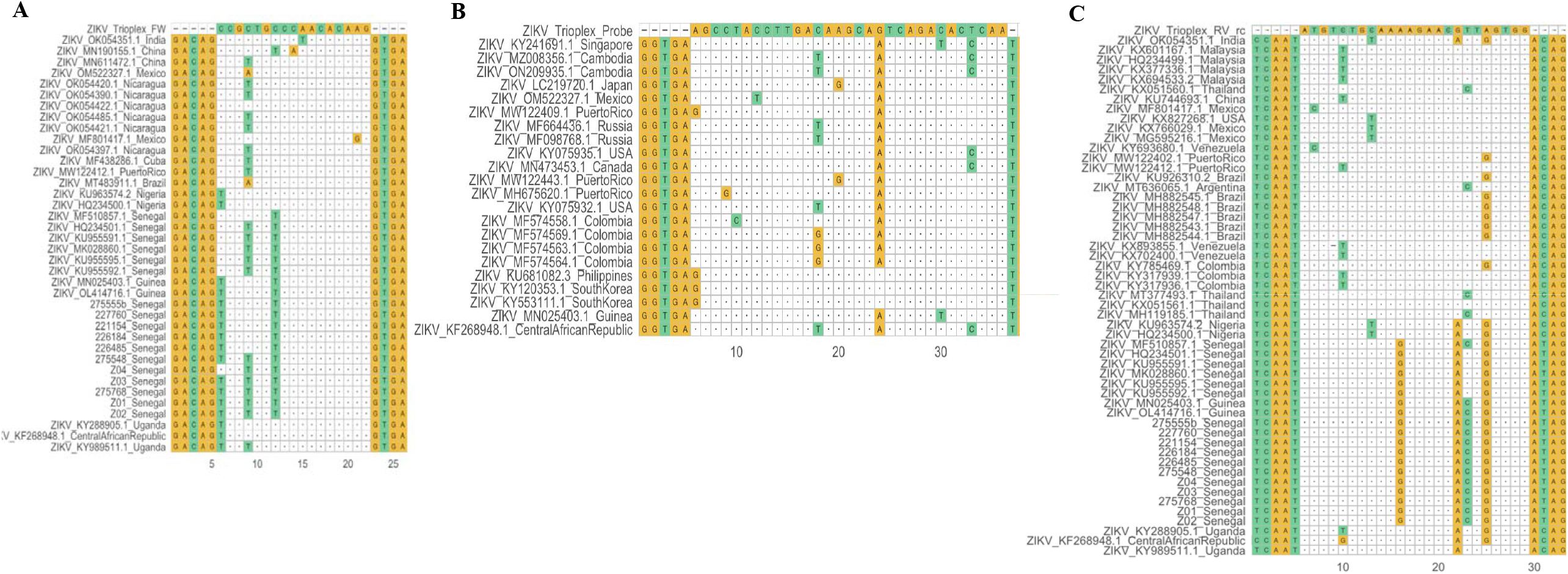
ZIKV CDC Trioplex oligonucleotide against representative sequences downloaded from Genbank. For each assay’s oligo only sequences exhibiting at least on mismatch was represented. A, B, and C indicate forward (FP), probe (P), and reverse (RP) oligonucleotide, respectively.

For certain West African strains in particular, we identified two to three mismatches within the first seven nucleotides of the forward primer’s 5’ extremity. While mismatches at the 5’ end of primers have been shown to have variable (and often minimal) effects on RT-qPCR performance (29,32), one Mexican strain exhibited a notable mismatch within the last two nucleotides of the forward primer’s critical 3’ extremity. In the reverse primer, the West African strains also demonstrated three to four mismatches, with three specifically located within the last eight, seven, and five nucleotides at the 3’ extremity. Mismatches near the 3’ end of primers are well-documented to significantly affect qPCR efficiency and often lead to higher cycle threshold (Ct) values (33–36).

The ZIKV CDC Trioplex probe also exhibited several mismatches, which are known to exacerbate RT-qPCR quantification errors (25–27). Within the forward primer, most mismatches were found between pyrimidine bases, though this balance shifts when considering the probe region. In contrast, the reverse primer presented mismatches upstream, within the core, and downstream of the sequence, predominantly involving purine-purine mismatches. These purine-purine mismatches are recognized to induce greater Ct value deviations compared to pyrimidine-pyrimidine mismatches (28,29). Additionally, several strains exhibited simultaneous mismatches in both primers, a condition widely associated with diminished RT-qPCR performance (37). This confluence of mismatches, particularly in key regions of the primers and probes, underscores the need for careful validation of the CDC Trioplex assay when used with these divergent viral strains.

## Conclusion

In conclusion, our study highlights the presence of multiple mismatches in the CDC ZIKV primers and probes across various strains circulating globally, with a notable prevalence in West African strains. Such mismatches, affecting both primers and probes, are well-documented to hinder the efficiency of RT-qPCR assays. Given the pivotal role of the CDC Trioplex assay in the simultaneous laboratory diagnosis of DENV, CHIKV, and ZIKV— offering numerous operational advantages—we recommend that laboratories, particularly in Western Africa, proceed with greater caution and perform prior validation of the CDC trioplex assay before routinely using it. However, further detailed studies are necessary to fully assess the practical implications of these mismatches on the Trioplex system’s diagnostic accuracy for ZIKV, especially in regions with diverse viral strains. Future research should include conducting experimental validations of primer-template mismatches to assess their impact on diagnostic efficiency, expanding genotype analyses to encompass a wider variety of ZIKV strains, and implementing field studies during outbreaks to evaluate the assay effectiveness in real-world settings. Additionally, there is a need to develop improved assays that can accommodate known variations in viral sequences, perform comparative studies with other diagnostic tools to identify their strengths and weaknesses in sensitivity and specificity, and focus on user training to standardize protocols for consistent results.

## Data Availability

All data produced in the present work are contained in the manuscript

## Author contributions

**Conceptualization**: Idrissa Dieng; **Data curation**; Idrissa Dieng; **Methodology:** Idrissa Dieng, Mignane Ndiaye, Diamilatou Balde,, Agathe Shella Efire, Manfred Weidmann; **Project administration:** Idrissa Dieng, **Resources:** Idrissa Dieng; Manfred Weidmann; **Software:** Idrissa Dieng; **Supervision:** Idrissa Dieng, Manfred Weidmann, Oumar Faye; **Validation:** Idrissa Dieng, Mignane Ndiaye, Diamilatou Balde, Agathe Shella Efire, Marielton Dos Pasos Cunha, Andrea Silverj; **Visualization:** Idrissa Dieng; **Writing – original draft:** Idrissa Dieng, Manfred Weidmann; **Writing – review & editing:** Idrissa Dieng, Mignane Ndiaye, Diamilatou Balde, Agathe Shella Efire, Marielton Dos Pasos Cunha, Andrea Silverj, Manfred Weidmann

