## Supplementary Files for "Global In-Silico Performance Assessment of the CDC Zika Trioplex Detection System"

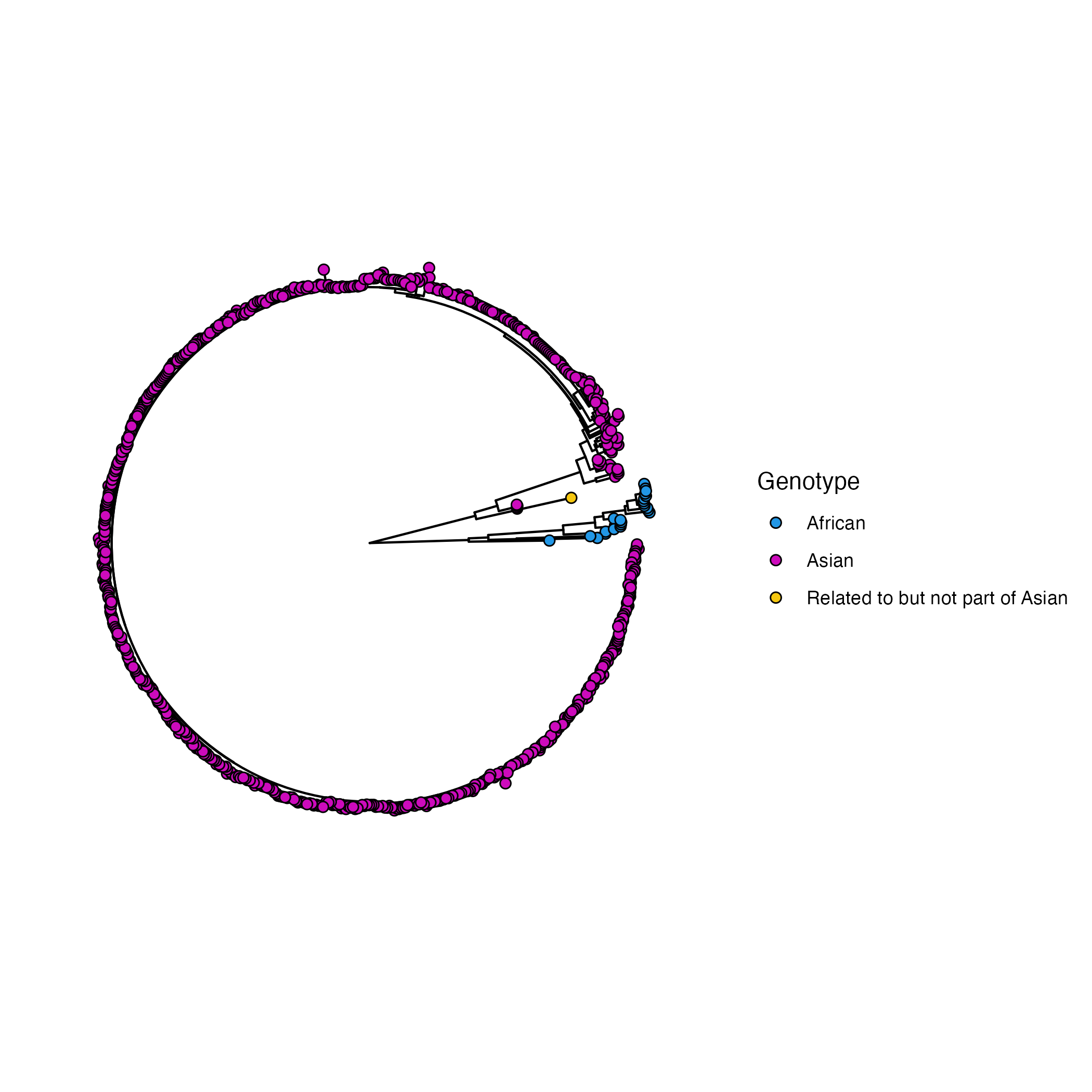

**Figure S1:** Maximum Likelihood (ML) tree showing the phylogenetic relationship of analyzed global ZIKV strains retrieved from Genbank. Sequences span the time period from 1966 to 2021. Sequences tips are colored according to ZIKV genotypes depicted using the Genomedetective ZIKA typing tool accessed at 18July 2024. An intermediate clade (yellow) is observed between dominant clades for Asian (violet) and African (blue) genome sequences.

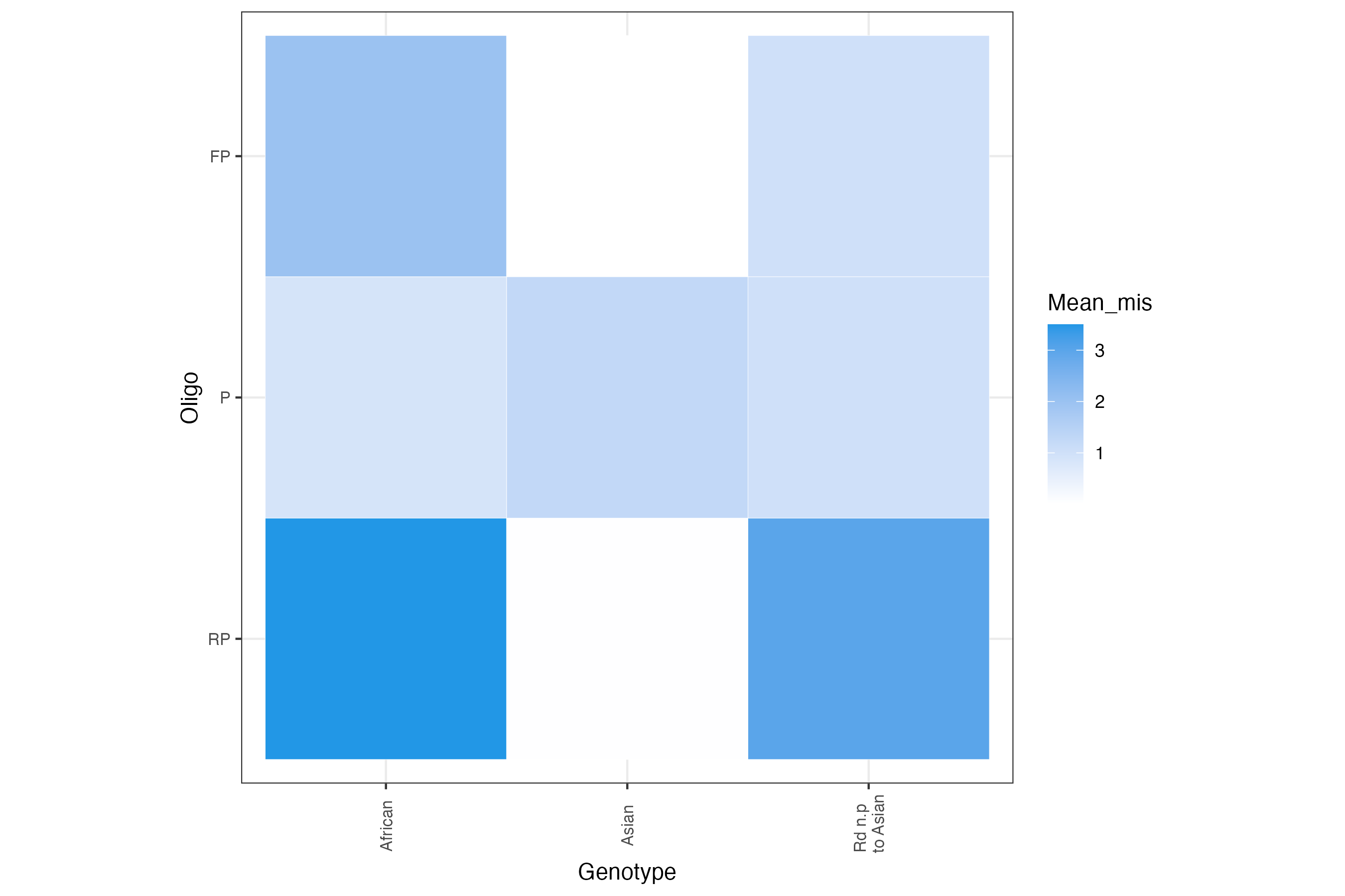

**Figure S2:** Mean oligonucleotides mismatches rates per assigned ZIKV genotypes using Genome detective.

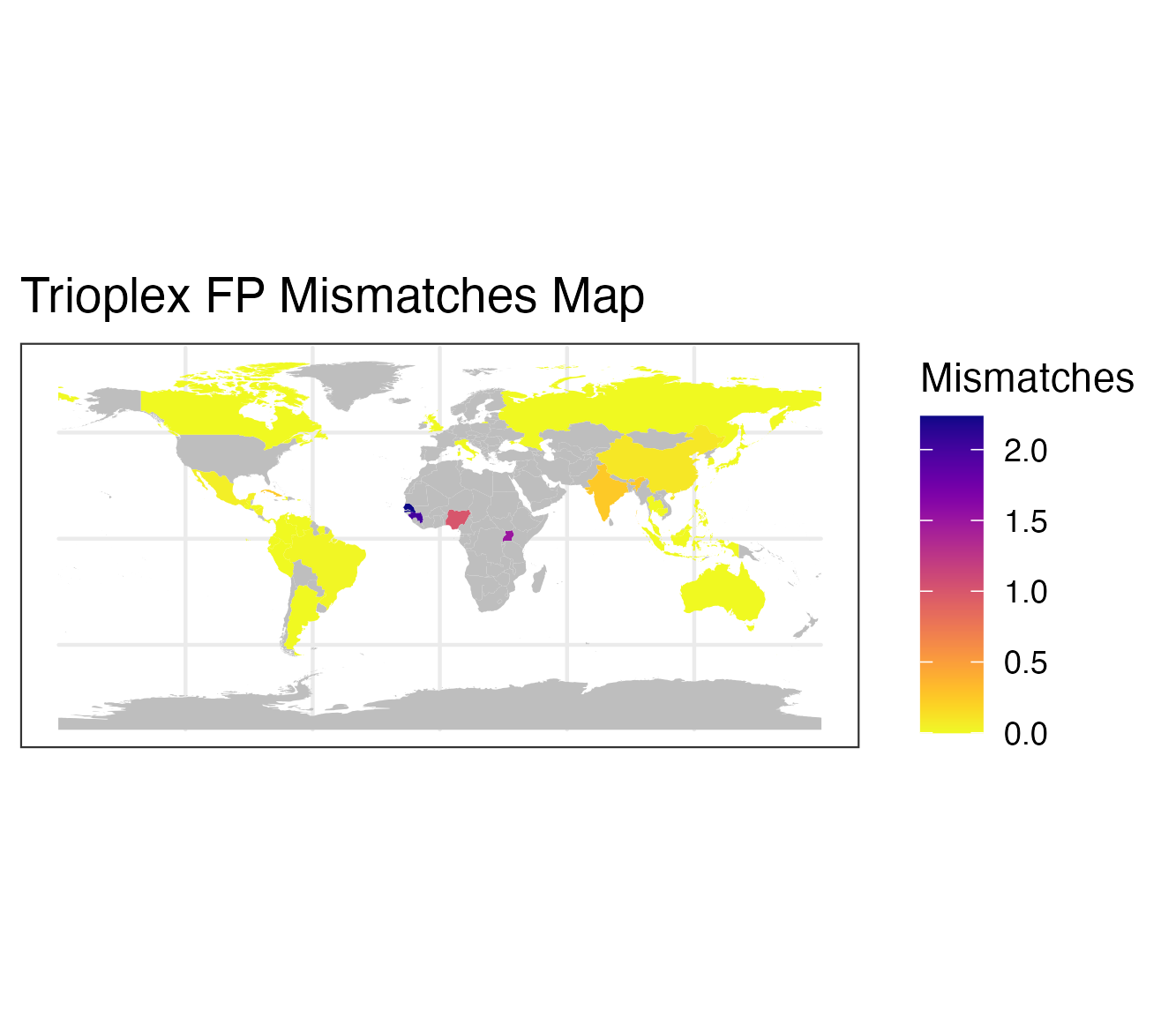

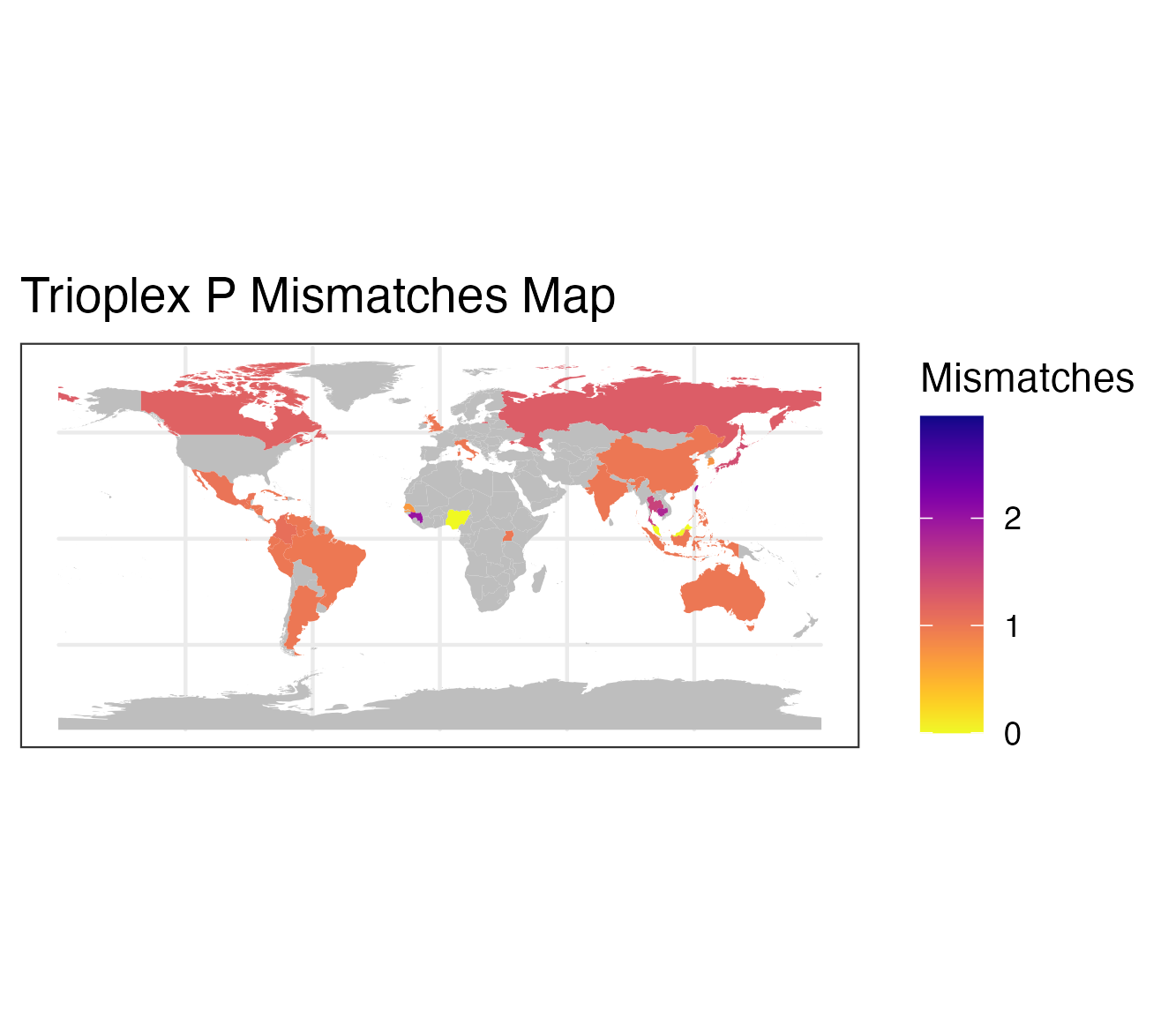

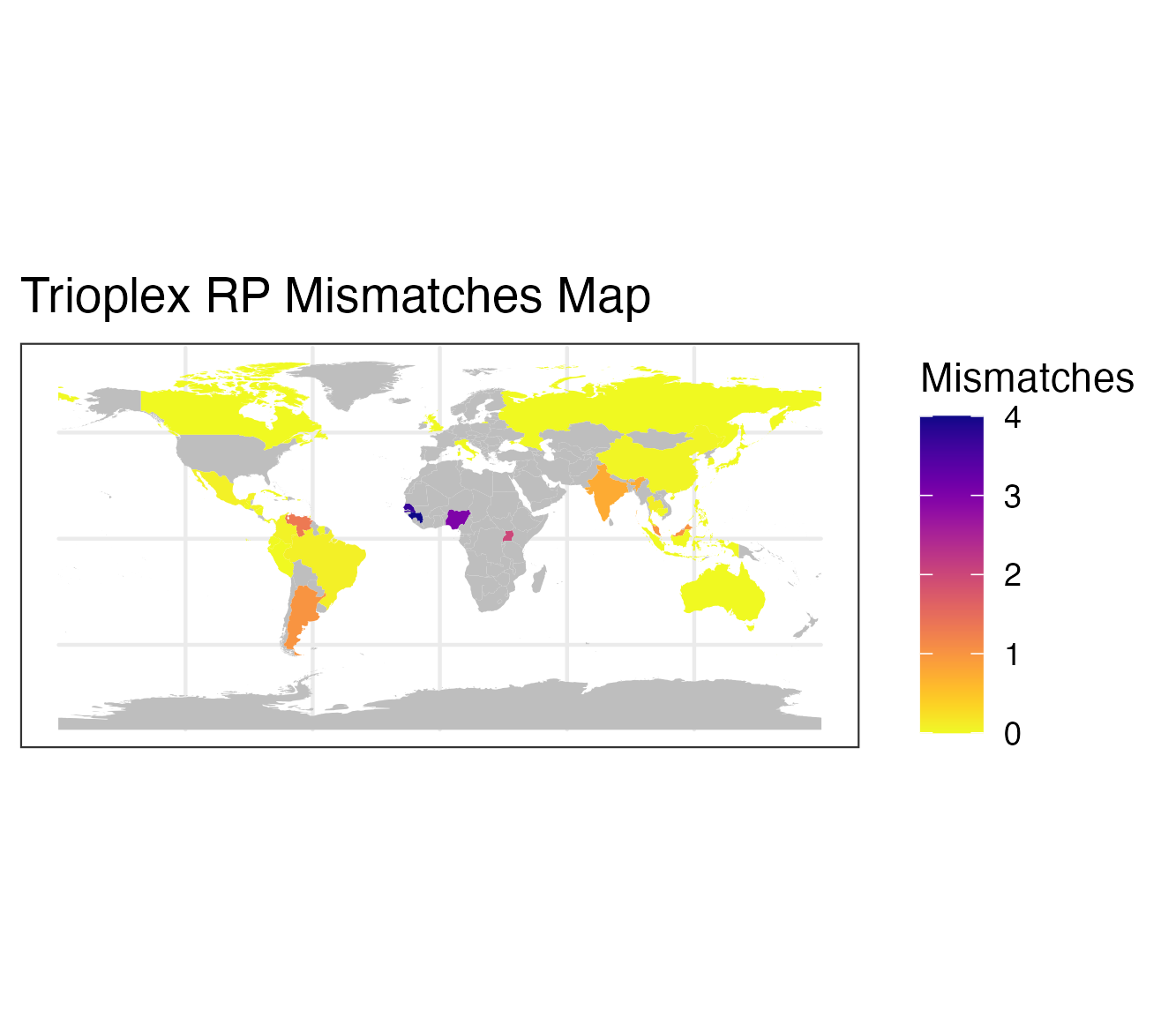

**Figure S3.** Country wise mean mismatches rate between ZIKV CDC trioplex assay oligos and representative dataset of viral lineages sequences. Countries in grey have no data
